# Change in stroke presentations during COVID-19 pandemic in South-Western Sydney

**DOI:** 10.1101/2022.05.02.22274456

**Authors:** James Thomas, David Manser, Peter Thomas, Nicholas Moore, Paul Middleton, Dennis Cordato, Cecilia Cappelen-Smith, Alan McDougall

**Affiliations:** Department of Neurophysiology, Liverpool Hospital, Liverpool, Australia; Department of Emergency Medicine, Liverpool Hospital, Liverpool, Australia; South Western Sydney Clinical School, University of New South Wales, Liverpool, Australia

## Abstract

**Background:** Australia managed relatively well during the global COVID-19 pandemic owing to our swift mandated public health response. During the NSW lockdown restrictions, we noted a decrease in acute stroke presentations at our institution, similar to what was subsequently reported worldwide.

**Aims:** We aimed to test our hypothesis that (i) the true numbers of ischaemic strokes did not change, however patients were presenting later and (ii) the proportion of TIAs decreased.

**Methods:** We conducted a retrospective audit of all stroke and TIA presentations in 2020 and compared these with data from 2019. We collected information about stroke subtype, severity, time from stroke/TIA onset to presentation and acute reperfusion therapies.

**Results:** Between January-February and April-March 2020, there was a 15% drop in acute stroke presentations (128 vs. 109). In the same period “stroke mimic” presentations dropped by 22%. The proportion of patients attending the emergency department within 4.5hrs was only 36% compared with 48% over the similar period in 2019.

**Conclusions:** Although the raw numbers of ischemic stroke presentations remained stable during NSW Covid lockdown, the proportion of patients presenting within time window for acute reperfusion therapies fell. The number of TIAs similarly fell suggesting COVID-19 discouraged patients from presenting to hospital which placed them at higher risk of disabling stroke. The opportunity cost of lockdown restrictions on stroke outcome should be considered in future policy directives.

## Introduction

Stroke is the third highest cause of mortality in the Australian population and the tenth leading specific cause of disease burden. Ischaemic stroke, caused by occlusion of cerebral vasculature, accounts for approximately 80% of stroke presentations(1). In 2017-2018 there were approximately 40,000 acute care hospital admissions for stroke, at a rate of 133 per 100,000 of population(2). The healthcare expenditure attributed to stroke is approximately $600 million per year. Estimates considering the long-term disability cost of stroke are up to $54 billion per annum(3).

One of the most important factors in determining outcome following ischaemic stroke is timely medical assessment and access to therapies such as intravenous thrombolysis (IVT) or endovascular thrombectomy (EVT). The IST-3 trial showed that patients presenting to a primary or comprehensive stroke centre within 4.5hrs of ischaemic stroke onset benefit from intravenous thrombolysis(4). Further trials using advanced imaging techniques for patient selection have shown a potential benefit of intravenous thrombolysis up to 9hrs from ischaemic stroke onset however the benefit diminishes with increasing duration between onset to treatment(5,6). Patients with ischaemic stroke due to large vessel occlusion benefit from EVT if performed within 6 hours(7,8). A subset of patients may benefit from treatment up to 24hrs (9–11). A number of strategies have been employed to minimise delays from symptom onset to reperfusion including but not limited to; public education initiatives to improve community awareness of stroke symptoms, pre-hospital triage and co-ordination, emergency department pathways and dedicated stroke teams.Access to inpatient stroke unit care, with specialist allied health providers and neurology trained nursing staff has been shown to further improve outcomes following acute stroke(12).

While timely treatment of stroke and inpatient stroke unit care have both been proven to improve outcomes following acute stroke, prevention of stroke is still far superior in preventing long term disability. Identification and treatment of individuals at high risk of stroke is a cornerstone of stroke prevention strategies. Transient ischaemic attacks (TIA) and/or minor strokes are well described to carry high risk of recurrent stroke (13). Timely identification of these events along with investigation and treatment of underlying risk factors has s significant effect at reducing recurrent stroke(14).

During 2020, the COVID-19 pandemic had a significant impact on the global population. Both internationally, as well as in Australia, lockdown restrictions were enacted to reduce community spread of the virus. These included restrictions on visiting family members, unnecessary travel and encouraged working from home for non-essential workers. There was significant media coverage of disease outbreaks and a high level of community awareness.

There have been a number of reports demonstrating an association between COVID-19 infection and acute ischaemic stroke, particularly in younger patients (15–17) though this has not necessarily correlated with an overall increased incidence of stroke.

Worldwide, particularly in countries that experienced a far higher burden on COVID-19 than Australia, there were reports of a significant change in the rates of acute stroke presentations to stroke centres. Several centres in the USA and Canada recorded a marked decrease in acute ischaemic stroke presentations (18–23). This trend was also seen in the UK, Italy, Norway and, to a lesser extent, in almost all countries with a developed stroke healthcare system (24–27). A similar trend was seen in Victoria, Australia, with a decrease in numbers of acute stroke presentations to the emergency department (28). While a decrease in acute stroke presentations or “code stroke” activations was universally reported, the rates of severe strokes, large vessel occlusions and reperfusion therapies offered varied significantly between sites. The majority of sites noticed a similar trend, that of reduced overall numbers of acute stroke presentations with a relative increase in the rates of ischaemic stroke due to large vessel occlusion (LVO), while the rates of minor stroke and TIA were reduced (18–20,22–27,29).

Several studies demonstrated a trend of increased time from onset of symptoms to ED presentation during the height of pandemic cases (18,29,30) though this data was not the case in a large cohort study from London (24), nor was this data reported in the Australian cohort.

In Liverpool Hospital, a comprehensive stroke centre in South-Western Sydney, we noted an apparent change in the numbers of stroke and TIA presentations through our emergency department, shortly after the announcement of lockdown measures in NSW.

## Aims

We aimed to test the hypotheses: (i) that the numbers of acute ischaemic stroke presentations would remain constant while the proportion of mild strokes and TIAs would fall, (ii) that patients with acute ischaemic stroke would present later than they had done pre-pandemic.

## Methods

We performed a retrospective analysis of all patients presenting to Liverpool Hospital with sudden onset neurological symptoms for whom the acute stroke team was activated in the Emergency Department (ED).

All patients presenting to Liverpool hospital ED with new onset neurological deficits are assessed in triage using the BE-FAST screening tool (31,32). Any patient with new onset balance issues; visual loss or diplopia; face, arm or leg weakness; and/or sensory change; either transient or persisting; with onset within the preceding 24hrs trigger the activation of a code-stroke for the acute stroke team. Demographic, imaging, treatment and time metric data are collected prospectively into a code stroke registry. We retrospectively reviewed all cases in the stroke registry and correlated this with the electronic medical record to obtain final diagnosis and discharge destination. To account for patients with milder symptoms or transient deficits who may not have been identified for acute stroke team activation at triage, we also included cases admitted under the neurology service with a diagnosis of transient ischaemic attack. Final diagnosis was recorded as either (i) acute stroke, including ischaemic and haemorrhagic strokes; (ii) transient ischaemic attack (TIA) or (iii) non-stroke neurological diagnosis, such as migraine, disorder of peripheral nerve, intracranial space occupying lesion, central nervous system inflammatory disorders and functional neurological disorders.

Liverpool Hospital, where the data was collected, is a metropolitan tertiary referral hospital and comprehensive stroke centre servicing a population of greater than 900,000 people. In addition to direct stroke presentations, we act as a 24/7 referral centre for endovascular clot retrieval as one of 3 centres within metropolitan Sydney. The local population is diverse with a large culturally and linguistically diverse population. Approximately 36% of the population were born overseas and 50% speak a language other than English at home (33).

We compared stroke presentations from 2020 with historical data from 2019 and earlier in attempt to control for natural variation in stroke presentations over the year. The first case of COVID-19 in Australia was identified on the 25th of January 2020 and the first case of community spread were identified on 2nd March 2020. Social distancing restrictions and closure of non-essential businesses were first enacted in NSW on the 21st of March 2020. After cases subsided in June 2020, a “second wave”, originating in the South-Western Sydney region began in early July 2020. A confirmed transmission event occurred in our emergency department in August 2020.

## Results

There were 847 acute stroke and/or TIA presentations in 2019 compared with 886 in 2020. The mean age was similar between years, 67.9 years in 2019 and 68.0 years in 2020. In 2019, 29% of patients were female compared to 47% in 2020. Between January-February and March-April 2020, there was a 15% reduction in stroke presentations (from n=128 to n=109). Stroke presentations rebounded by 77% to 193 in May-June 2020. (Fig. 1)

**Figure 1.**
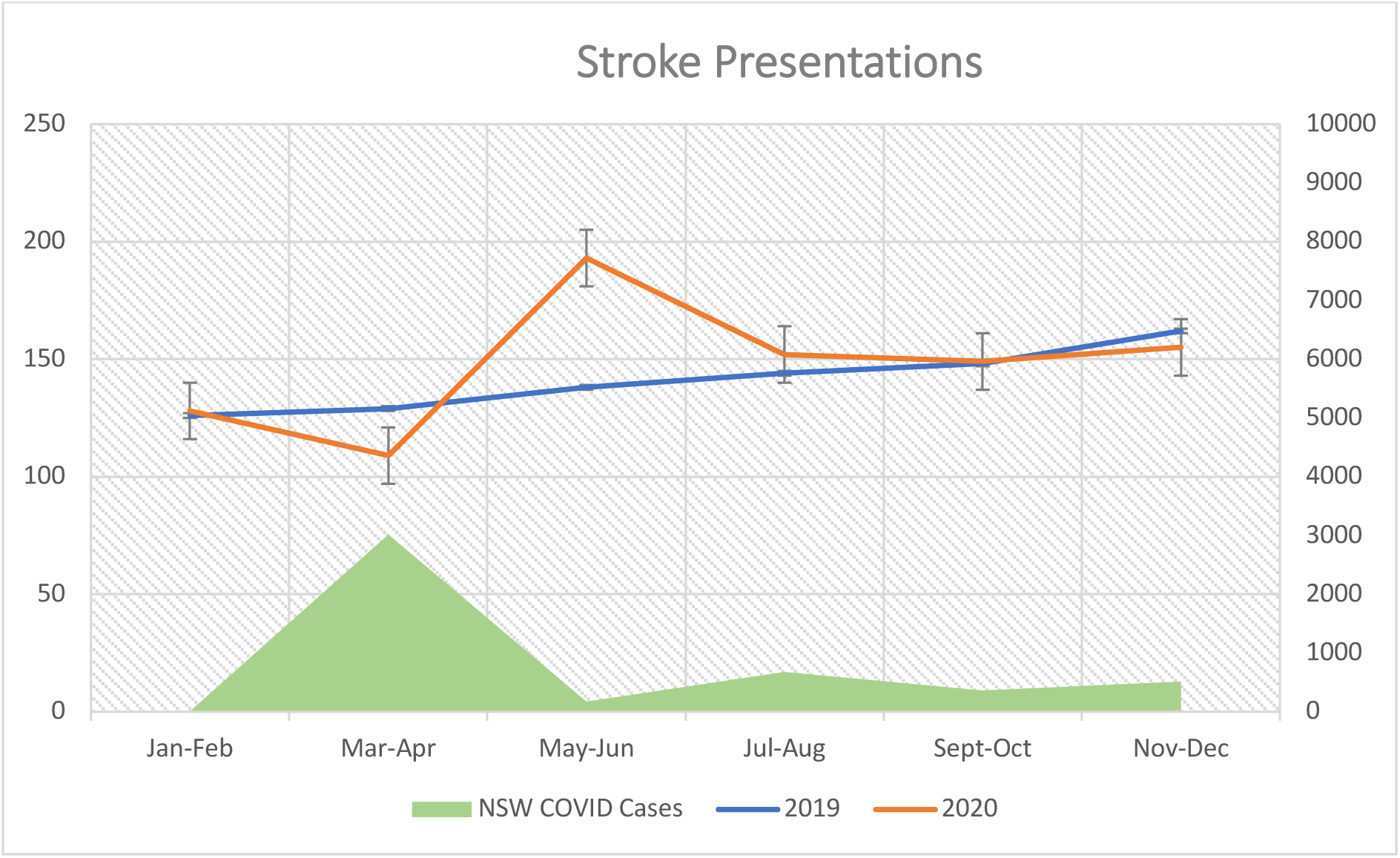
Total stroke presentations vs. NSW reported COVID-19 cases. Error bars = standard deviation of monthly stroke cases

We reviewed the medical record for all patients who activated a code-stroke on arrival to ED and extracted their final discharge diagnosis. When classified by final diagnosis, the most significant difference in presentations was seen within the patients with a final diagnosis other than stroke or TIA. Examples of non-stroke activations include Bell’s palsy, syncope, delirium, seizures and post-ictal states, migraine, and functional neurological disorders. (Fig.3) Between January-February and March-April, “non-stroke” stroke presentations fell by 22%. In May-June, the number of “non-stroke” presentations rose by 104%. TIA presentations saw a reduction from 15 to 10 (−33%) with a rebound to 27 (+170%) while strokes remained stable between January-February and March-April 53 to 52 (−2%) before increasing to 70 (+38%). (Fig. 2 & Fig. 4)

**Figure 2.**
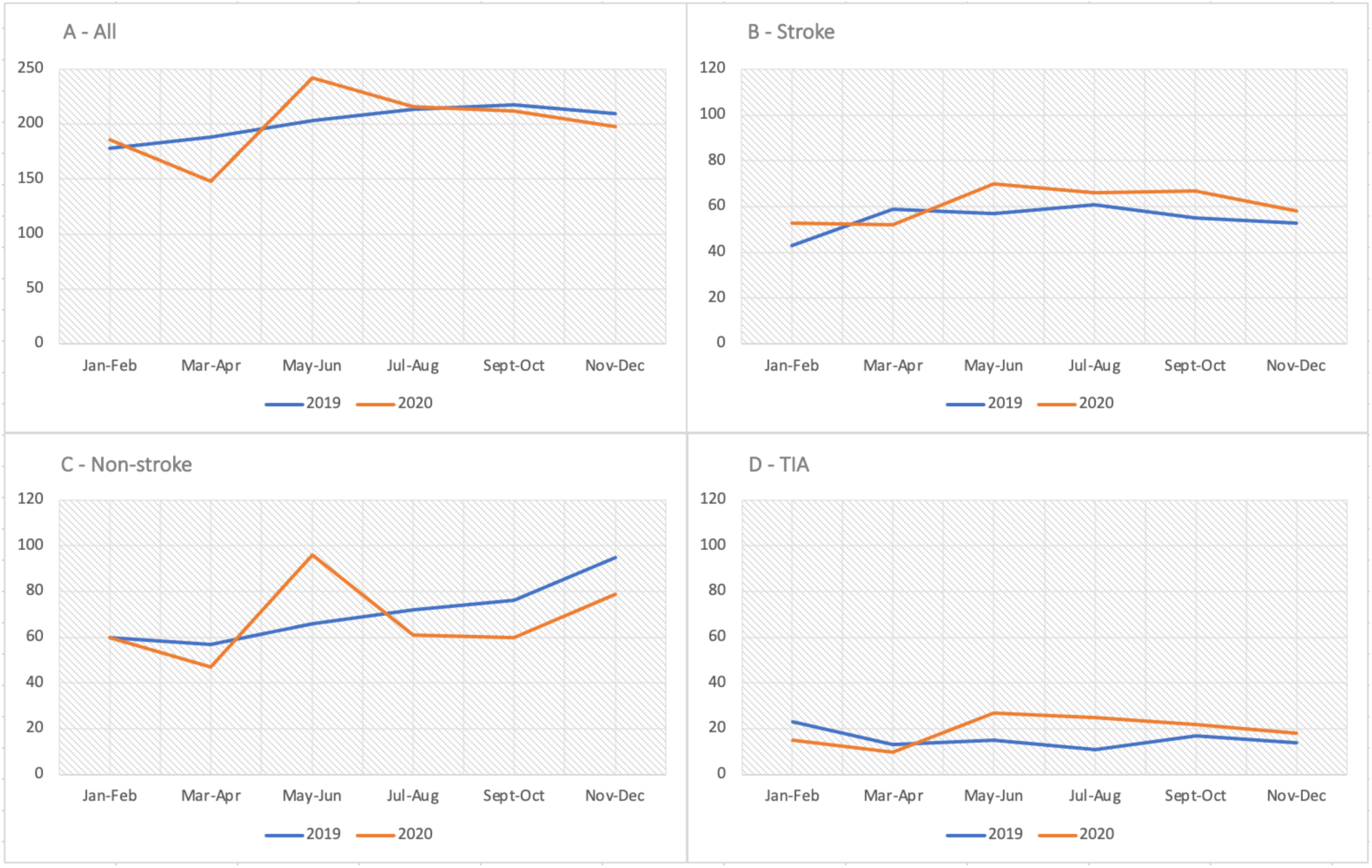
“Code-Stroke” case numbers. a) total acute stroke presentations, b) final discharge diagnosis of stroke (ischaemic or haemorrhagic), c) final diagnosis of other neurological condition, d) final diagnosis of transient ischaemic attack

**Figure 3.**
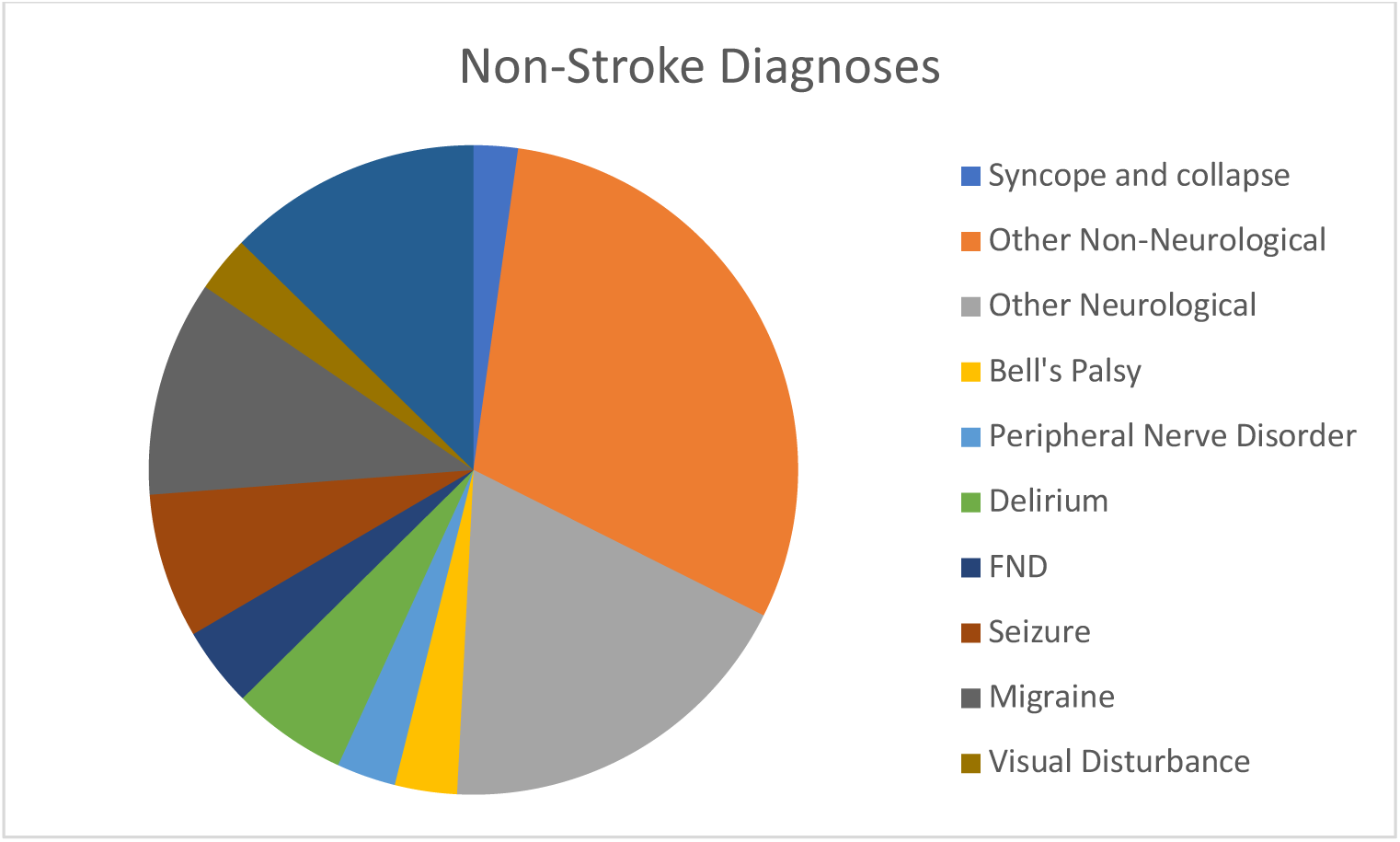
Final Diagnosis of Stroke Presentations other than stroke or TIA. (Non-stroke)

**Figure 4.**
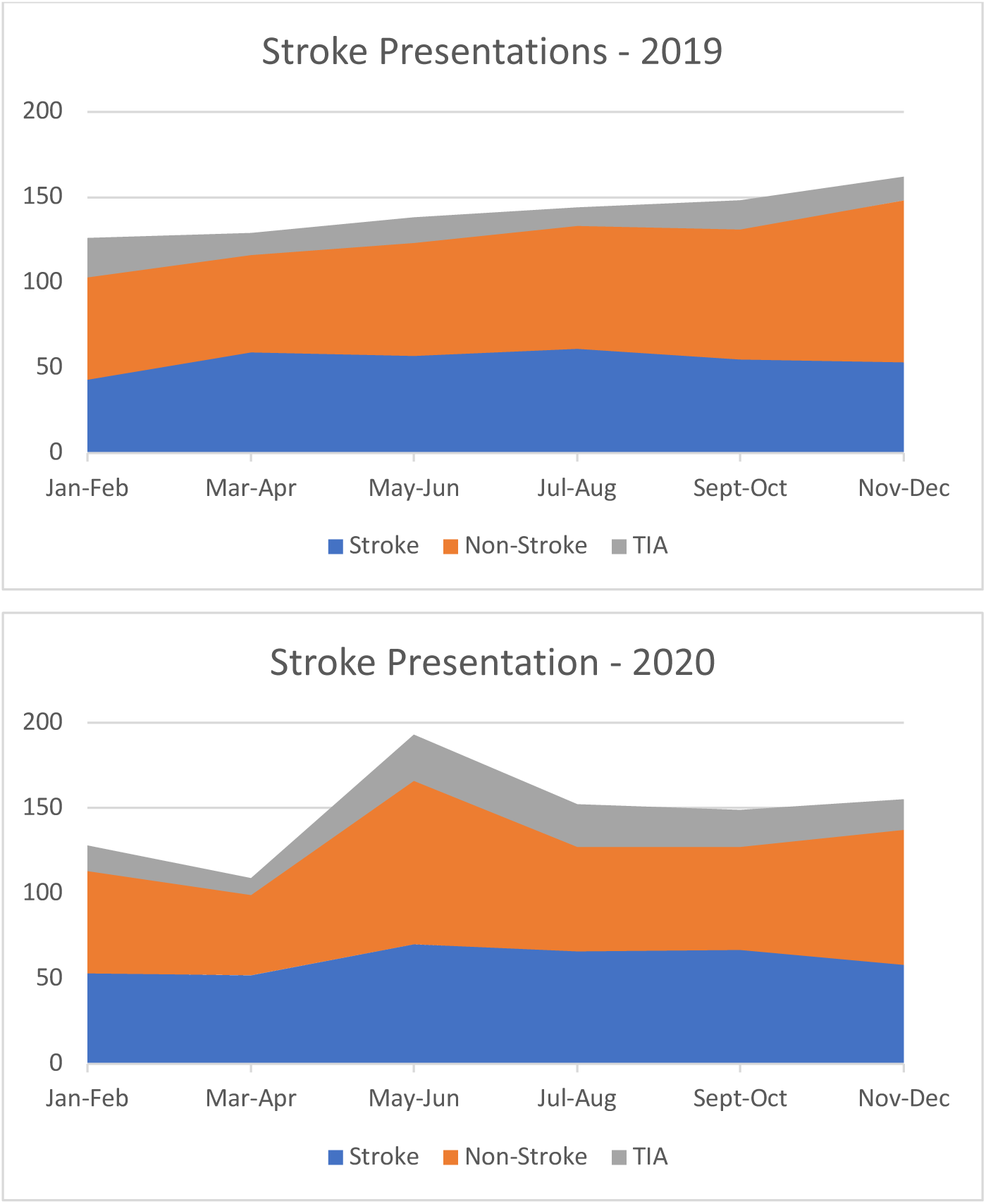
Bimonthly breakdown of “Code-Stroke” activations by final diagnosis in 2019 and 2020

The proportion of patients presenting within 4.5hrs of symptom onset in March-April fell from 48% in 2019 to 36% in 2020. This rebounded to 44% in the period May-June 2020, a similar proportion to 2019. (Fig. 5)

**Figure 5.**
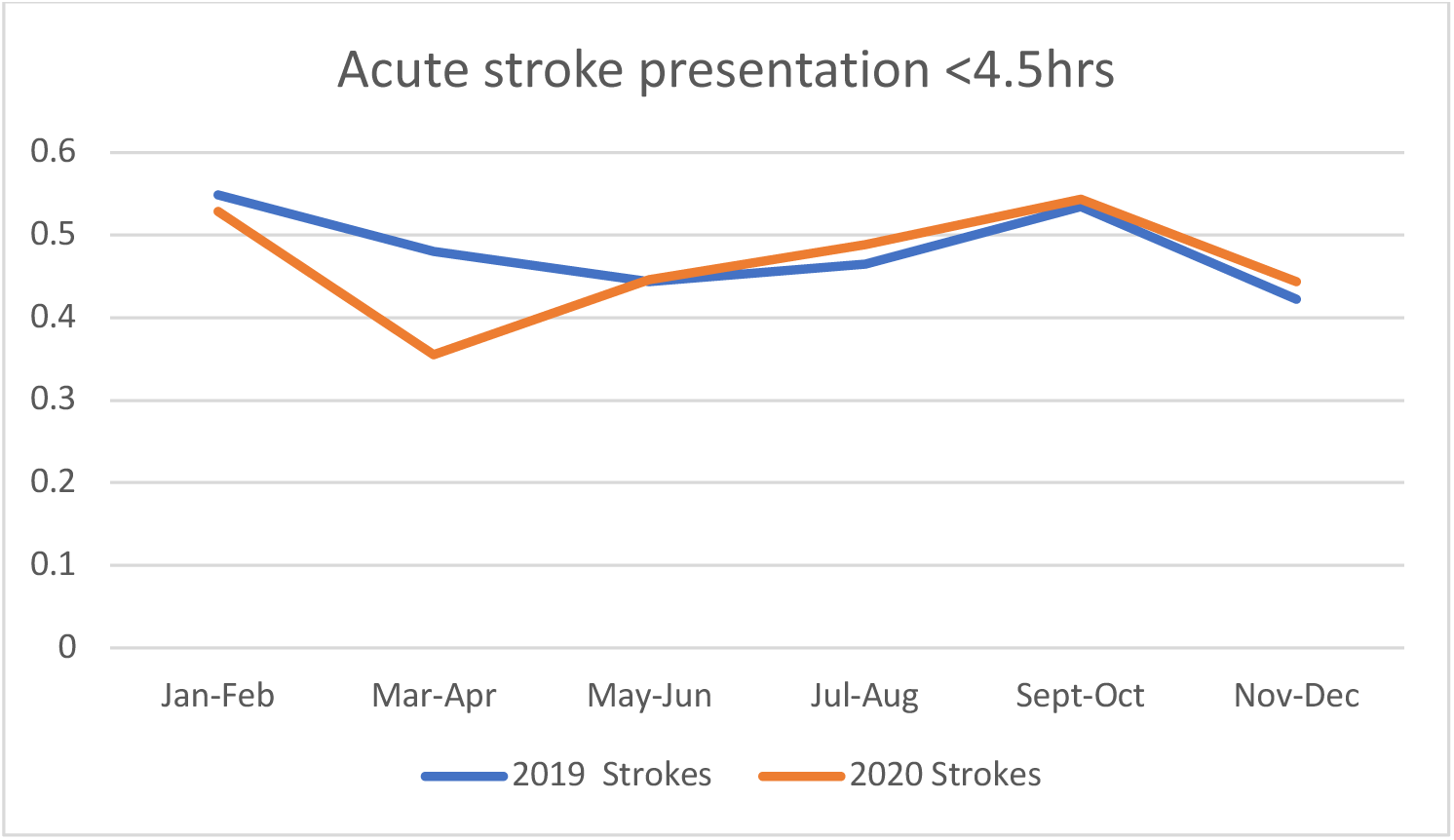
Bimonthly proportion of strokes presenting within 4.5hrs of symptom onset in 2019 and 2020.

The mean National Institute of Health Stroke Score (NIHSS) was 10 for both years. In April 2020, the mean NIHSS increased to 12 compared with 10 in April 2019. The mean NIHSS peaked at 13 in July 2020 compared with 11 in July 2019. (Fig. 6)

**Figure 6.**
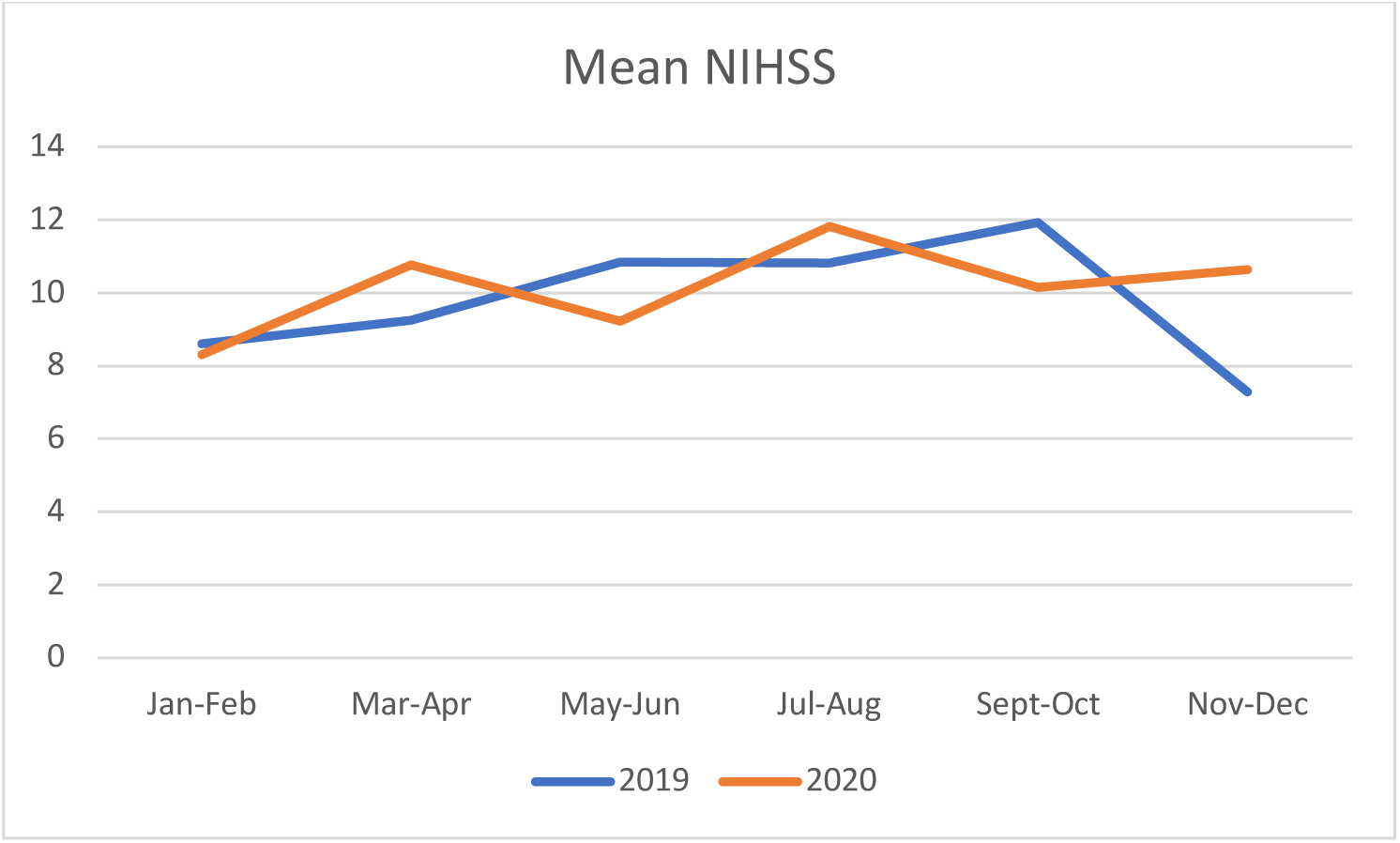
Bimonthly mean NIH Stroke Scale for 2019 and 2020

The average delay from presentation to intravenous thrombolysis was 71 minutes in 2019 and 77 minutes in 2020. There was similar month to month variability in both years. A similar trend was seen in delay from door to arterial puncture in those eligible for endovascular clot retrieval with an average delay of 146 minutes in 2019 compared to 124 minutes in 2020. In 2020, 13% of all acute stroke presentations received either intravenous thrombolysis and/or endovascular clot retrieval, compared with 11% in 2019. The average length of stay was similar between years at 5 days.

## Discussion

During the first wave of the COVID-19 pandemic many countries reported a dramatic decrease in acute stroke presentations. This change appeared to be driven, in part, by a decrease in numbers of mild strokes and/or TIA presenting to hospital. The cause for this reduction in stroke presentations remains unclear, particularly given evidence suggesting a causative relationship between acute COVID-19 infection and acute stroke. Australia had fewer cases of COVID-19 infection per capita compared with most other developed countries reporting this trend. We identified a similar pattern of stroke presentation in a region without significant strain on the health infrastructure by high numbers of COVID-19 infections.

In our centre, we retrospectively collated data from several electronic record sources to describe the pattern of acute stroke presentations before and after the onset of the first wave of the COVID-19 pandemic in New South Wales. We noted a modest decrease of acute stroke presentations during the two-month period March-April while COVID-19 cases were increasing in the community, and public health restrictions were being enacted. This decrease in acute presentations was driven by a reduction in presentations of acute neurological deficits that did not have a final diagnosis of ischaemic or haemorrhagic stroke. Presentations of TIA and stroke also fell, albeit to a lesser degree. The initial decrease in stroke presentations was followed by a rebound of 77% in the period May-June. The initial decrease in cases was like that reported in the literature (15,17–22,24–28,34–39). We saw a reduction in stroke presentations despite a relatively low incidence of SARS-CoV-2 infections compared to other centres internationally. This finding provides support for the assertion made by several authors (18,20,30,35,36,40,41) that the reduction in stroke presentations may be related to patient reluctance to seek medical attention for symptoms for fear of contracting COVID-19 infection rather than external barriers to accessing health services such as hospitals and emergency medical services overwhelming their capacity.

The significant rebound following the lockdown period lifting was an unexpected finding and not one reported elsewhere. A concern expressed in several publications was that a reduction in mild stroke or TIA presentations would place people at increased risk of subsequent severe stroke as they would not receive adequate evaluation and management of modifiable stroke risk factors (15–17,21,37,40–43). This seems less likely to be the case in our cohort as the reduction in mild stroke and TIA presentations were far fewer than the excess in presentations in the post-lockdown period. The increase in stroke presentations following lockdown seen at our institution was also driven largely by an increase in non-stroke diagnoses. Stroke severity, as measured by mean NIHSS also did not differ significantly between pre and post lockdown periods.

Another of our findings was that the proportion of patients presenting within 4.5 hours of stroke onset fell significantly in the period of lockdown. This has great clinical significance as the period up to 4.5-hour post onset of stroke, sometimes referred to as the “golden hour”, represents the time in which acute reperfusion therapies such as intravenous thrombolysis have class 1 evidence for reducing long term disability from acute ischaemic stroke (44). Significant investment in patient education, pre-hospital infrastructure and ED protocols has been made to reduced delays in diagnosis and management of acute ischaemic stroke to increase access to reperfusion therapies. This finding of a decrease in stroke presentations within 4.5 hours may be attributed to several factors. As postulated earlier, concern about being exposed to COVID-19 may initially deter patients from seeking medical attention. Even if patients do seek attention, there may be an unnecessary delay between onset of symptoms and medical contact. Initial lockdown orders in NSW discouraged any movement outside the home, including visiting family members. Some stroke syndromes, particularly those involving the non-dominant cerebral hemisphere may have a significant element of anosognosia. These presentations rely heavily on family members identifying the symptoms of stroke and prompting evaluation. Many other stroke symptoms, such as hemiplegia or aphasia produce significant barriers in seeking medical attention without family or caregiver support. To our knowledge, there were no significant procedural related delays relating to pre-hospital care and transport.

Our study was limited by its retrospective observational design at a single centre which prevents robust conclusions being able to be drawn about causation of this change to stroke presentations. Code-stroke activations, the basis for including patients in our analysis, are initiated after initial clinician assessment in the ED and are subject to some degree of bias towards more serious presentations. That is, patients with mild or transient symptoms may not trigger an initial code-stroke activation. We attempted to improve our capture of such patients by also reviewing all patients admitted under the neurology service. It is the policy at our institution that all patients with minor stroke and/or TIA are admitted for initial workup rather than triaged to an outpatient service. Another limitation of our study is that it was underpowered to identify significant differences in the stroke subgroups due to low numbers of strokes in a relatively short period of lockdown.

New South Wales is currently in the middle of a period of significant increase in COVID-19 transmission due to a local outbreak of the delta strain of the virus in Western and South Western Sydney. We plan to repeat our analysis on the new time period in order to increase our statistical power strengthen our results.

## Conclusion

The pattern of acute stroke presentations in South Western Sydney, during a period of highly effective community lockdown restrictions highlights the potential downsides of strict lockdown regulations on non-COVID-19 health outcomes. While the initial lockdown measures in NSW were highly effective in rapidly reducing community spread of COVID-19, this may have been at the expense of inadvertently discouraging patients from seeking medical attention for time-critical conditions such as stroke. Future consideration should be taken to ensure patients are willing and able to access medical care in a timely manner, regardless of the situation in the community.

## Data Availability

All data produced in the present study are available upon reasonable request to the authors

